# Scalable Transdiagnostic Early Assessment of Mental health (STREAM): Study Protocol

**DOI:** 10.1101/2024.05.07.24306697

**Authors:** Elin H. Williams, Nicholas. M. Thompson, Gareth McCray, Maria. M. Crespo-Llado, Supriya Bhavnani, Diksha Gajria, Debarati Mukherjee, Teresa Del Bianco, Georgia Lockwood-Estrin, Luke Mason, Vukiwe Ngoma, Chisomo Namathanga, Richard Nkhata, Allan Bennie, Alok Ranjan, Ulemu Kawelama, Naina Midha, Anindita Singh, Innocent Mpakiza, Akshat Gautam, Sheffali Gulati, Mark H. Johnson, Gillian Lancaster, Matthew K. Belmonte, Emily J. H. Jones, Vikram Patel, Sharat Chandran, Emmie Mbale, Gauri Divan, Melissa Gladstone, Bhismadev Chakrabarti

**Author notes:** to whom correspondence should be addressed Professor Bhismadev Chakrabarti School of Psychology and Clinical Language Sciences, University of Reading, Reading, RG6 6ES.

## Abstract

**Introduction:** Early childhood development forms the foundations for functioning later in life. Thus, the ability to accurately monitor developmental trajectories is critical. However, such monitoring is complicated by reliance on time-intensive assessments which often necessitate administration by skilled professionals. This difficulty is exacerbated in low-resource settings where such professionals are predominantly concentrated in urban and often private clinics, rendering them inaccessible to a large portion of the population. This geographic and economic inaccessibility contributes to a significant ‘*detection gap*’ where many children who might benefit from additional support remain undetected. The Scalable Transdiagnostic Early Assessment of Mental Health (STREAM) project aims to bridge this gap by developing an open-source, scalable, tablet-based mobile platform administered by non-specialist workers to assess motor, social, and cognitive developmental status. The ultimate goal is to deploy STREAM through public health initiatives, maximising opportunities for effective early interventions.

**Methods and analysis:** Four thousand children aged 0-6 years from Malawi (n=2000) and India (n=2000) will be enrolled and assessed using the STREAM platform. STREAM integrates three established developmental assessment tools measuring motor, social, and cognitive functioning using gamified tasks, observation checklists, parent-report, and audio-video recordings. Domain scores for motor, social, and cognitive functioning will be developed and assessed for their validity and reliability. These domain scores will then be used to construct age-adjusted developmental reference curves.

**Ethics and dissemination:** The STREAM project has received ethical approval from local ethics review boards at each study site, is carried out in accordance with Good Clinical Practice (GCP) standards, and adheres to ethical guidelines presented in the 6th (2008) Declaration of Helsinki.

**Strengths and limitations of this study:**

- This study will collect data from N=4000 children in Malawi and India to assess the reliability and validity of the Scalable Transdiagnostic Early Assessment of Mental Health (STREAM) tablet-based platform;
- Community-based recruitment and the inclusion of children with known or suspected neurodevelopmental conditions will enable us to test the utility of STREAM in identifying children whose developmental status is below that expected for their age;
- The user-friendly nature of the tablet-based platform, coupled with its data security features and adaptability to different linguistic and cultural contexts, enhances its potential for widespread use in different populations;
- The reference curves generated in this study are expected to be applicable in the sampled contexts but may necessitate validation in other settings;
- The duration of the STREAM assessment in its current version may pose challenges, and is an area for refinement following data analysis.

## INTRODUCTION

Optimal development and good mental health in early childhood form the foundations for positive outcomes in later life, such as improved retention in education, better employment prospects, and an overall higher quality of life (Black and Hurley, 2014). Accordingly, the United Nations (UN) Sustainable Development Goals (SDGs) Target 4.2 highlights the critical need for all children to have access to quality early life care to ensure they have the best chance of achieving their developmental potential (UN SDGs, 2015). Accomplishing this goal necessitates the availability of appropriate tools for assessing children’s development to identify those with functional difficulties (Richter et al., 2019; Waldman et al., 2021). Such tools can facilitate the implementation of effective interventions in early childhood, when brains are maximally plastic and responsive to changes (Patel et al., 2018).

Many tools designed to assess difficulties in early childhood functioning face significant limitations. A majority of these tools focus on identifying or diagnosing specific neurodevelopmental conditions such as Autism Spectrum Conditions (ASC) or Intellectual Disability (ID), rather than assessing neurodevelopmental status using a dimensional framework, similar to those proposed by the Research Domain Criteria (RDoC; Insel et al., 2010) and the Hierarchical Taxonomy of Psychopathology (HiTOP; Kotov et al., 2017). Some tools that do measure development dimensionally, such as the Guide for Monitoring Child Development (GMCD; Ertem et al., 2008), rely only on parent-report and/or clinician-observation measures, which can be affected by recall bias, subjectivity, and inter-rater variability. Critically, most tools are only applicable for a relatively narrow window within the early developmental period. For example, the recently developed Global Scales for Early Development (GSED; McCray et al., 2023) and Caregiver Reported Early Development Instruments (CREDI; McCoy et al., 2016, 2018) target only the interval from birth to three years of age. In contrast, the Save the Children’s International Development and Early Learning Assessment is appropriate for children aged 3 to 6 years (Halpin et al., 2019). Although some tools enable assessment throughout the early developmental period (e.g., Griffiths Mental Development Scales [GMDS; Griffiths, 1996], Ages and Stages Questionnaires [ASQ; Bricker & Squires, 1999]), these are hindered by issues of scalability: They are time-intensive, proprietary and incur significant financial costs for training and implementation, requiring administration by skilled professionals. Further, the majority have been developed to high-income country norms, limiting their applicability in other contexts (Gladstone et al., 2010; Peña, 2007; Waldman et al., 2021).

These limitations of existing tools highlight a need for a tool that assesses development dimensionally, measuring functioning across multiple domains and throughout the years of early development. This need is particularly pressing in low-resource settings where access to skilled professionals may be limited. Low and middle-income countries (LMICs) face significant resource constraints (Dasgupta et al., 2016; Durkin et al., 2015) and children in these settings are disproportionately exposed to risk factors known to impact development, including poverty, violence, inadequate hygiene and cognitive stimulation, perinatal issues, and poor nutrition (Bitta et al., 2017; Toso et al., 2020; Byrne et al., 2017; Shonkoff et al., 2012). Such risk factors can adversely affect both physical and mental health, hinder cognitive development, and contribute to poor long-term outcomes (Bhutta et al., 2023; Shonkoff et al., 2012). These factors may partly explain why an estimated 50.2 million children in LMICs meet the criteria for some form of neurodevelopmental condition (Olusanya et al., 2018) and an estimated 250 million children stand at risk of not meeting their developmental potential (Grantham-McGregor et al., 2007).

The Scalable Transdiagnostic Early Assessment of Mental health (STREAM) project aims to overcome the various limitations of existing developmental assessment tools. STREAM is a digital, tablet-based platform that assesses motor, social, and cognitive functioning in children aged 0 to 6 years. Our objectives for the STREAM platform are that it is: 1) able to assess motor, social, and cognitive abilities across the early developmental period; 2) applicable across diverse cultural settings; and 3) scalable and usable by non-specialist workers (NSWs).

We will construct normed reference curves for each developmental domain measured (motor, social, cognitive). The broader, long-term use of such reference curves is to track and identify children with atypical developmental trajectories. By enabling early identification of such children, these reference curves will facilitate timely referrals and appropriate early intervention.

### Aims

1. To develop a tablet-based tool to measure motor, social, and cognitive abilities of children aged 0-6 years in two low-resource settings;
2. To generate normative reference curves of motor, social, and cognitive abilities;
3. To establish criterion validity of the domain scores (motor, social, cognitive) against an established measure of child development (GMDS; Griffiths, 1996);
4. To establish the convergent validity of the STREAM platform by assessing the relationship between domain scores and known correlates of development via self-report, biological, and neural measures;
5. To establish the test-retest reliability of the scores;
6. To establish the responsiveness of the scores (sensitivity to change) against changes in the GMDS, in a longitudinal assessment of a subsample of children.

## METHODS

### Design and study sites

The STREAM project, set in India and Malawi, is a cross-sectional study with an additional longitudinal component. Malawi is categorised as one of the world’s least developed countries, while India falls within the lower-middle income category, as defined by the Organisation for Economic Cooperation and Development (OECD, 2023). These two countries vary in terms of language, culture, and medical/educational infrastructure, which enables the assessment of the STREAM platform’s potential generalisability across diverse contexts.

### Patient and public involvement

The feasibility and acceptability to stakeholders of the three established tools included within the STREAM tablet-based platform have been assessed previously (Gladstone et al., 2010; Dubey et al., 2024; Bhavnani et al., 2019). Local stakeholders within the recruitment catchment areas were consulted before commencing STREAM data collection. In India, Accredited Social Health Activists (ASHA) provided feedback on our proposed referral pathways for children and families requiring more specialist support (see Referrals and Support section). In Malawi, feedback on recruitment, acceptability, feasibility, as well as strategies for strengthening local capacity was provided by paediatricians, the Association of Early Child Development, District Health Officers, Community Health Workers, and local schools. Additionally, participating families provided feedback on assessment burden and feasibility of the project during piloting.

### Study Sample

The study sample will comprise 4000 children aged between 0-6 years. Children will be recruited to either a *Community* (N=3700) or *Enriched* sample (N=300). The *Community* sample will consist of children recruited from Blantyre, Malawi, and New Delhi, India (specific participant recruitment protocols and selection criteria are outlined in a subsequent section). The *Enriched* sample will include children from tertiary clinical centres diagnosed with or showing characteristics indicating a high likelihood of having a neurodevelopmental condition (e.g., ASC). By recruiting these children, we aim to provide a proof of principle for the reference curves generated from the STREAM platform by testing children whose developmental status is known to be below that expected for a typically developing child of their age.

### Recruitment and Consent

A quota-sampling approach will be implemented to ensure adequate representation across sex and age categories (Supplementary Materials 1 and 2). To monitor recruitment progress, we will conduct quarterly reviews starting from the commencement of data collection. If, during these quarterly reviews, certain age and sex categories are underrepresented in the recruited sample, targeted recruitment efforts will be undertaken.

### Community Sample

A database of potential participants will be established through liaison with governmental health service providers (ASHA in India and Health Surveillance Assistants in Malawi) for parents, or expecting parents, in antenatal, immunisation, and weighing clinics operating within the catchment areas. Families will be approached, either at clinics, at home, or by telephone, and informed of the objectives and assessment procedures of the study. Subsequent recruitment will be achieved through snowball sampling via word of mouth. Interested families will be provided with information sheets and screened for eligibility (detailed below) after providing informed consent. Only one child from each household will be eligible to participate.

In India, children (N=1850) aged 0 to 6 years will be recruited from the urban South-East District of New Delhi. In Malawi, children (N=1850) aged 0 to 6 years will be recruited from Limbe and Ndirande Health Centres, Blantyre District, where families typically access routine healthcare services such as vaccinations. While the majority of children attending these health centres for routine appointments in Malawi will likely be younger children (under three years), we will extend invitations to parents who also have other children who are older than three years. Children older than three may also be recruited from local primary schools and Early Childhood Development centres.

### Community Sample Inclusion and Exclusion Criteria

Children will be eligible for STREAM if:

1. They are between 0 and 72 months of age (i.e., 0-6 years);
2. Their parent/caregiver can provide informed consent;
3. They and their parent/caregiver reside within the catchment areas of the study sites.

Children will be excluded if:

1. Their sibling has participated in the STREAM study;
2. They have a severe vision, hearing, or motor impairment, as reported by their parent/caregiver, which would limit their ability to interact with a tablet device;
3. They have had an uncontrolled seizure in the last 48 hours that lasted more than 5 minutes;
4. They are currently enrolled in another research study or trial;
5. Their parent/caregiver has a severe vision or hearing impairment;
6. Their parent/caregiver has a severe learning disability or a current, severe psychiatric condition.

### Enriched Sample

In India, children diagnosed with a neurodevelopmental condition (N=150) will be recruited through liaison with tertiary hospitals (e.g., All India Institute of Medical Science; AIIMS). These children will have a pre-existing diagnosis from an experienced clinician using the Diagnostic and Statistical Manual of Mental Disorders (5th Ed; DSM-V).

Recruitment of children for the *Enriched* sample in Malawi (N=150) will target various healthcare facilities, including paediatric wards, paediatric neurology clinics, physiotherapy clinics, occupational therapy clinics, psychiatric clinics, and malaria follow-up clinics, at the Queen Elizabeth Central Hospital (QECH). Additionally, children will be recruited from centres for children with special needs including the Hamilton’s centre, Feed the Children, and Jacaranda. Given resource limitations in Malawi, we anticipate that many children will not have pre-existing diagnoses of neurodevelopmental conditions. However, their attendance at these specialised clinics and centres reflects their greater likelihood of exhibiting characteristics indicative of having a neurodevelopmental condition (e.g., motor or cognitive difficulties). Clinical officers will assess whether each recruited child meets the inclusion criteria for the *Enriched* sample (detailed below) after checking their health passport and/or medical files and observing their behaviour.

In light of variations in the typical age of onset and detection for different neurodevelopmental conditions, recruitment to the *Enriched* sample at both sites will target different phenotypes across age strata (n.b. a formal diagnosis of any neurodevelopmental condition is not necessary for inclusion in the *Enriched* sample). For instance, our recruitment strategy will target children aged 0 to 2 years who are either diagnosed with Global Developmental Delay (GDD) or are exhibiting characteristics of GDD. For children aged 2 to 4 years, we will target those diagnosed with GDD, Autism Spectrum Conditions (ASC), or Intellectual Disability (ID), or exhibiting characteristics of these conditions. Lastly, for children aged 4 to 6 years, we will target those diagnosed with ASC, ID, or ADHD, or exhibiting characteristics of these conditions. While ADHD is typically diagnosed after 6 years of age (American Psychiatric Association, 2013), early signs and ‘red flags’ are often observed between 4 and 6 years.

### Enriched Sample Inclusion and Exclusion Criteria

In addition to the inclusion criteria^1^ outlined above for the *Community* sample, children recruited to the *Enriched* sample must meet one of the following criteria:

1. A pre-existing clinical diagnosis of a neurodevelopmental condition (e.g., GDD, ID, ASD, ADHD) or
2. Documented indications of developmental delays in their health passport or medical records, or such delays are observed by a clinical officer.

**Figure 1.**
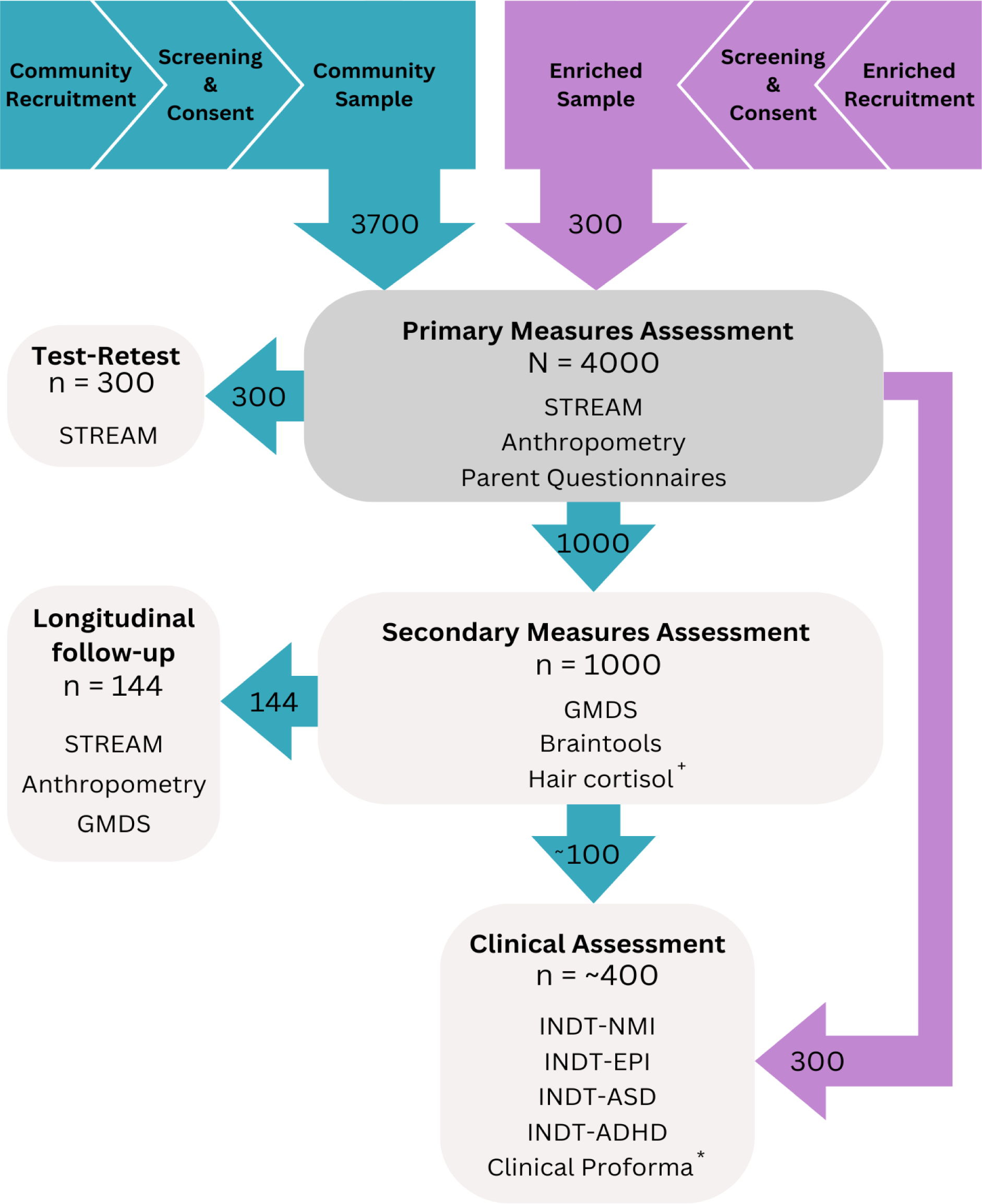
STREAM recruitment and assessment components. *As children recruited to the *Enriched* sample in Malawi are unlikely to have pre-existing diagnoses, they will additionally be administered a clinical proforma. ^+^ Hair samples will be collected from a smaller subsample of n=200.

### Primary Measures Assessment (PMA)

All (N = 4000) children from the *Community* and *Enriched* samples will be administered the PMA (Figure 1) which includes i) the STREAM platform (Figure 2), ii) anthropometric measures (i.e., height, weight, mid-upper arm circumference, head circumference), iii) parent/caregiver report of neurodevelopment through the Rashtriya Bal Swasthya Karyakram (RBSK; Singh et al., 2015), and iv) questionnaires measuring socio-demographics and exposure to risk factors relevant to neurodevelopment (details of all measures included in STREAM can be found in Supplementary Materials 3).

### STREAM Platform

The STREAM platform’s content and design are built upon three established and complementary child development measurement tools, previously developed, field-tested, and validated by the team for use in low-resource settings: the Malawi Developmental Assessment tool (MDAT; Gladstone et al., 2010), DEvelopmental assessment on an E-Platform (DEEP; Bhavnani et al., 2019), and the Screening Tool for Autism Risk using Technology (START; Dubey et al., 2024). The integration of these tools within STREAM was piloted on N=15 children in India and N=17 in Malawi to assess the need for cultural adaptation and to inform the development of Standard Operating Protocols (SOPs) and assessor training procedures.

### MDAT

The MDAT combines observational and performance-based assessments with parent-reported checklists, covering the following domains: gross motor, fine motor, language, and social. MDAT has strong psychometric properties and serves as a reliable instrument for identifying children aged 0 to 6 years in low-resource settings with delayed development and/or neurodisability (Gladstone et al., 2010). Any culture-specific content within the MDAT was adapted in consultation with the tool developer, and thorough translations, back-translations, and piloting were conducted at both sites prior to their inclusion within the STREAM platform.

### DEEP

DEEP is an innovative tablet-based tool designed to assess a range of cognitive processes such as manual speed and coordination, inhibitory control, visual perception and integration, reasoning, categorisation, and memory in preschool children aged 2.5 to 6 years. It comprises 14 games that are integrated into an overarching storyline which aims to maximise a child’s attention and engagement with the tool. Metrics derived from children’s interaction with DEEP predict their performance on the cognitive domain of the Bayley Scales of Infant and Toddler Development (BSID-III, Bayley, 2006; Mukherjee et al 2020).

### START

START is a mobile, modular, open-source platform originally designed for early detection of autism risk in children aged 2.5 to 6 years. The START tool assesses several domains associated with the autistic phenotype, including social functioning, sensory preference, and fine motor skills. It uses tablet-based tasks, incorporating performance-based metrics, video-based eye-tracking, and a video recording of parent-child interaction (PCI). It has shown high accuracy (>86%) in classifying children with a neurodevelopmental condition (ASC or ID) in field settings (Dubey et al., 2023). The START stimuli have been adapted for STREAM to ensure cultural and linguistic suitability at both sites. Additionally, any language content has been translated, back-translated, and piloted at both sites.

Each of these three tools will generate multiple output measures in STREAM (e.g. a single motor function assay will generate measures of spatial and temporal error). These output measures will be combined to derive the STREAM domain scores (i.e. social, cognitive, motor) (see *Data Analysis* section for specific details).

All children will be administered the MDAT observation checklists, and the START PCI component and social vs. non-social preferential looking task. Children aged 2.5 to 6 years will additionally be administered the gamified tablet-based tasks from DEEP and START.

In order to test if a child’s performance on tablet-based assessments may be influenced by their prior exposure to smartphones or tablets, we will assess exposure through parent-report. We will allow time for children with less exposure to tablet/smartphone devices to practise using the tablet screen (e.g., taps, drag and drop) before administering the STREAM tablet-based platform. During this familiarisation phase, which will last approximately 10 minutes, children will engage with a (non-STREAM-related) game on the device, enabling them to learn how to interact with a touch-screen device.

**Figure 2.**
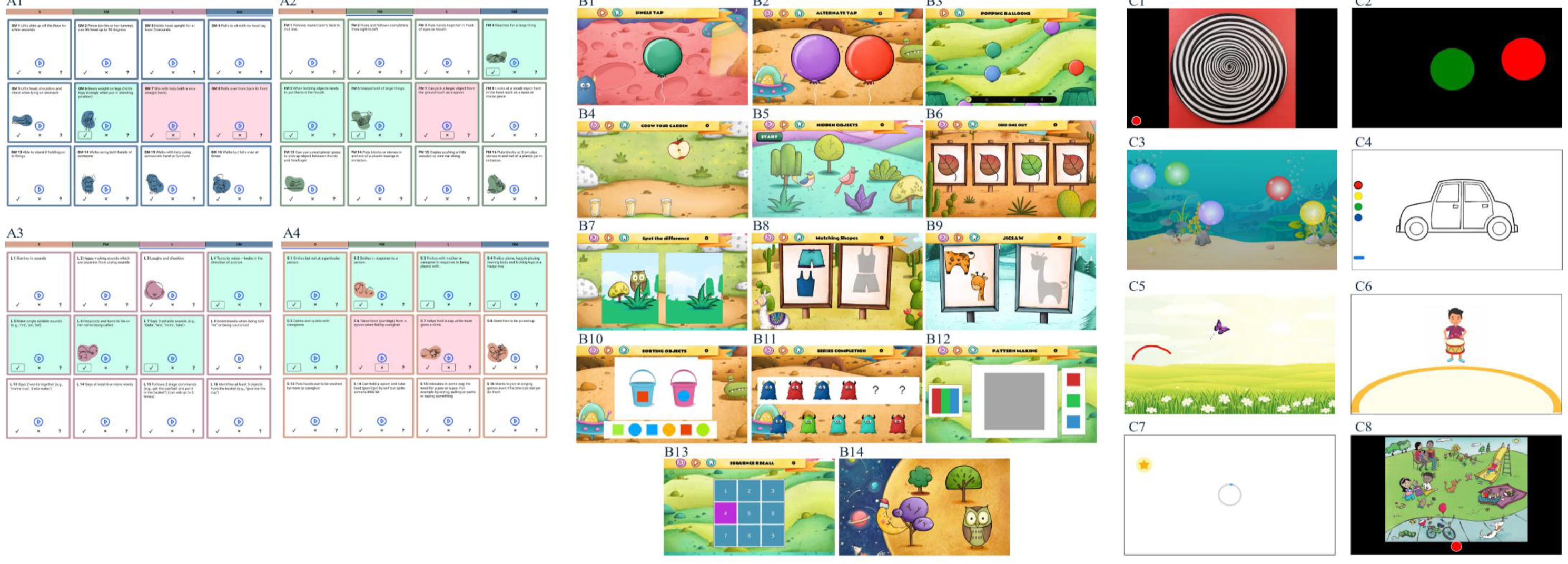
Visual depiction of each task within the STREAM platform. A: MDAT (a subset of items from each grid); A1: gross motor; A2: fine motor; A3: social; A4: language. B: DEEP; B1: single tap; B2: alternate tap; B3: popping balloons; B4: grow your garden; B5: hidden objects; B6: odd one out; B7: spot the difference; B8: matching shapes; B9: jigsaw; B10: sorting objects; B11: series completion; B12: pattern making; B13: sequence recall; B14: location recall. C: START; C1: wheel task; C2: button task; C3: bubble popping task; C4: colouring task; C5: motor following task; C6: synchrony task; C7: delayed gratification; C8: language sampling task. n.b. The images for the preferential looking task and the parent-child interaction are not included in this preprint version due to the publisher’s policy.

### Test-Retest (TR)

To test the STREAM platform’s TR reliability, a subsample of n=300 children (stratified by age and sex) will be randomly selected from the *Community* sample to be re-administered the STREAM assessment after an interval of 7-10 days.

### Secondary Measures Assessment (SMA)

One thousand children from the Community sample, stratified by age and sex (Supplementary Materials 4), will be administered i) the GMDS, and ii) concurrent eye-tracking and electroencephalogram (EEG) measures (i.e., Braintools). A hair sample will be collected from a smaller subsample (n=200) for the analysis of hair cortisol concentration. This SMA component will occur no later than 3-4 working days after the PMA.

### GMDS

The GMDS is widely used to measure child development across the 0 to 6 year age range. It provides a clinically assessed, continuous measure of strengths and needs in multiple domains (gross & fine motor coordination, language & communication, personal social-emotional function, and learning). A trained clinical psychologist or clinical officer at each site will assess each child. GMDS scores will provide dimensional measures of a child’s developmental attainment, against which the STREAM domain scores will be benchmarked as a measure of criterion validity. Although the GMDS was originally standardised in the United Kingdom, it has been widely used throughout the world (e.g., Laughton et al., 2010; Davies et al., 2011; Tso et al., 2018). To ensure cultural and linguistic suitability at both sites, we have adapted some items in consultation with the tool developers (n.b. details of this adaptation work will be reported in a separate manuscript).

### Braintools

The Braintools battery (Haartsen et al., 2021; Del Bianco et al., 2023a; Lockwood Estrin et al., 2022) will use EEG to examine brain responses to social, communicational, and sensory stimuli. Concurrent eye-tracking will be employed to achieve two objectives: 1) to implement gaze-contingent tasks, where trials progress when the child is attending appropriately to the stimuli presented on the computer screen, and 2) to capture visual attention to social and non-social stimuli (see Supplementary Materials 3 for a list of Braintools tasks). The social stimuli have been adapted for cultural and linguistic suitability at both sites. The measures obtained via this component will provide a test of convergent validity for the domain scores derived from the STREAM assessment. For example, we will examine the extent to which the STREAM social domain scores correlate with neural responses to social stimuli.

### Hair Cortisol

Hair samples of approximately 3 cm in length will be collected by trained assessors from n=200 children to measure hair cortisol concentration. Cortisol level in hair has been demonstrated to be a robust non-invasive indicator of stress in the preceding months and is associated with neurodevelopmental outcomes (Bhopal et al., 2019). This measure will be used to assess the convergent validity of the STREAM domain scores.

### Clinical Assessment (CA)

To understand the clinical characteristics of the *Enriched* sample at both sites, we will administer four INCLEN Diagnostic Tools (INDTs). All children aged 0 to 6 years in this sample will be administered the INDT for Epilepsy (INDT-EPI; Konanki et al., 2014) and INDT for Neuromotor Impairment (INDT-NMI; Gulati et al., 2014). Children older than 12 months will be administered INDT for Autism Spectrum Disorder (INDT-ASD; Juenja et al., 2014), and those older than 48 months will be administered INDT for Attention Deficit Hyperactivity Disorder (INDT-ADHD; Mukherjee et al., 2014). These tools will be administered by a trained clinical psychologist or clinical officer to either confirm the presence of a neurodevelopmental condition or characterise the nature of the functional difficulties experienced by the child. Thorough translations, back-translations, and piloting of these tools has been conducted at both sites to ensure linguistic and cultural appropriateness.

As we anticipate that children recruited to the *Enriched* sample in Malawi are unlikely to have pre-existing diagnoses that could aid in characterising their functional difficulties, a clinical officer will administer a clinical proforma (Supplementary Materials 5) alongside the INCLEN diagnostic tools. This proforma will provide additional insights into a child’s difficulties and enable expert judgement regarding the presence/absence of a neurodevelopmental condition and the specific nature of any difficulties experienced by the child.

Furthermore, a subsample of children from the *Community* sample who have undergone both the PMA and SMA will be screened for neurodevelopmental conditions. Children who exhibit difficulties in RBSK neuromotor, motor, cognitive, or social domains will be invited to participate in the CA. A flag indicating higher likelihood of a neurodevelopmental condition will be assigned to children scoring 1 in any of these domains on the RBSK. Based on typical neurodevelopmental condition population prevalence rates (Arora et al., 2018), we expect that approximately 10% of children from the *Community* sample who have completed the SMA (n=100) will be identified as being at higher likelihood of having a neurodevelopmental condition and will be invited for this screening.

### Longitudinal Follow-Up (LFU)

A subsample of children (n=144; stratified by age and sex) who complete the SMA will be followed up and re-administered the STREAM platform, GMDS, and anthropometric measures after an 18-month interval to assess the responsiveness (sensitivity to change) of STREAM scores compared to GMDS scores.

### Assessor Training

To ensure scalability, the PMA will be administered by NSWs who meet the qualifications recommended by the governments of India and Malawi for frontline health workers (i.e. aged between 25 to 45 years with a minimum of senior school education). NSWs will be trained by senior project personnel in administering all components of the PMA (i.e., STREAM platform, questionnaires, anthropometry). This training programme is scheduled over eight days and involves a combination of classroom training with some explanation of child development, practical demonstrations of tool administration, and role-playing exercises. Trainees will receive feedback from trainers as well as their peers. Following this initial training, NSWs will engage in field practice for approximately 4-8 weeks to ensure they demonstrate proficiency in administering each component of the PMA and will meet regularly with trainers to address concerns or issues. Refresher training sessions will be scheduled every three months during the entire data collection period, although refresher training might occur more frequently during the early stages of data collection.

The SMA and CA will be conducted by clinical psychologists or clinical officers. These assessors will be trained in administering the GMDS after completion of a two-part course provided by the Association for Research in Infant and Child Development (ARICD). They will be observed and evaluated by a certified GMDS trainer on at least two occasions. Braintools training will consist of multiple online tutorials complemented with a comprehensive SOP (Del Bianco et al., 2023b) to ensure consistent execution of the protocols. Trainees will initially practise administering Braintools on adults before progressing to assessing children. All data collected by trainees will be reviewed by a senior EEG technician. Feedback on data quality will be provided, along with guidance on areas for improvement. Training for collecting hair samples for the analysis of hair cortisol concentration will consist of a 1-day online tutorial. Following this training, assessors will have access to an SOP with step-by-step instructions for sample collection. Training on the administration of the four INDTs will consist of a 2-day online tutorial provided by AIIMS. The trainee’s progress during the first few weeks of INDT administration will be supervised by a senior paediatrician. Clinical officers or psychologists will be introduced to the clinical proforma by a senior paediatrician. The senior paediatrician will provide a demonstration on how to administer and code each item of the proforma. The trainee’s progress during the first few weeks of clinical proforma administration will be supervised by the senior paediatrician.

### Data Storage and Quality Control

Anonymised data collected offline on the STREAM platform using a tablet are encrypted and later uploaded to a secure central STREAM back-end server located in Mumbai, India. Questionnaire, anthropometry, SMA, and CA data will be stored on REDCap, a web-based system for data collection (Harris et al., 2009), which uses a different server located in Belgium. Only approved research staff will have the requisite credentials for accessing the STREAM back-end and REDCap database. Videos on the STREAM back-end will be viewable only after the user uploads a secret decryption key and agrees to terms and conditions for video data handling.

Each site has a dedicated data quality officer (DQO) responsible for verifying data collected on both STREAM and REDCap platforms. The DQO will flag any instances of missing or incomplete data and report any anomalies detected in the data, thereby ensuring that potential issues with data, administration, or deviations from SOPs can be addressed quickly. Bi-weekly meetings involving principal investigators, senior staff, postdocs, data staff, and site personnel will monitor the ongoing progress of the project and provide a platform to address any questions or concerns raised by site teams.

### Statistical Analysis

In order to optimise the information gathered from STREAM and derive three key neurodevelopmental domain scores (i.e., motor, social, cognition) and validate the platform, we will perform two largely separable analysis pathways: 1) a ‘construct-focused’ approach and 2) a machine-learning approach. These two approaches for generating the STREAM domain scores will be reviewed once both analyses have been completed. The decision as to which will constitute the primary scoring approach for STREAM will be taken at a later date and will be outlined in a subsequent paper.

### Construct-focused score creation

In *step 1* of the construct-focused method of score construction, we will collate judgements from six STREAM subject matter experts (SMEs) regarding which domain(s) each task/metric in STREAM assesses. In *step 2,* all output metrics from each task will be constructed using the relevant guidelines for that tool. In *step 3,* three factor analysis/structural equation^2^ models (Kline, 2023) will be created and compared on data from n=1850 randomly subsampled children, with a n=1850 hold-out sample. Comparing the fits of the factor models will tell us whether the domain scores are statistically distinct and whether a *total* score is justifiable. Model 1, the baseline comparator, will be unidimensional with all items loading on one factor. Model 2 will use the SME judgement data to ascribe one task to one domain (i.e., no cross-loading). Model 3 will allow cross loadings in cases where the judgement data indicate that a task is measuring more than one domain. Model 4 *will not* allow cross loadings and will also include an additional second-order factor representing a “total score”. Model 5 *will* allow cross loadings and will also include an additional second-order factor representing a “total score”. Once each model is fitted, all metrics loading less than 0.34 on any factor will be removed iteratively. The inferences about the final scoring models will be based on interpretation of absolute (Chisq, RMSEA, CFI) and comparative (AIC, BIC, ADJ-BIC) model fit, using standard cut values (Dunn & McCray, 2020). We will decide whether: 1) a total score is to be modelled, and 2) cross-loading of tasks should be allowed. The final selected model will be fitted on the hold-out sample and the fit statistics will form evidence for structural validity. Then, the chosen model will be fitted on all data (N = 3700) and the final model parameters will provide the scoring model. In line with WHO child growth standards (WHO, 2006), we will construct reference curves for age-adjusted development scores. The planned reliability and validity analyses are outlined in Table 1.

**Table 1.** Overview of planned reliability and validity analysis.

| Evidence type | Sample size | Measure(s) | Analysis | Hypotheses |
| --- | --- | --- | --- | --- |
| Structural Validity | 1850 (MDAT only)<br>1234 (all tools) | STREAM scores | SEMs fitted on a random n=1850, stratified by age and sex, with n=1850 hold out sample for validation | RMSEA < 0.06<br>CFI > 0.95<br>Chi-sq |
| Internal consistency | 4000 | STREAM scores | Cronbach's Alpha (imputation of missing by design data) | Alpha > 0.90*<br>(each scale) |
| Test-retest Reliability | 150 | STREAM scores | Intra-class correlation coefficient (2,1) | ICC(2,1) > 0.75* |
| Criterion Validity (concurrent) | 1000 | GMDS Scores | Pearson's Correlation | R<sub>p</sub> > 0.90*<br>(Construct focused method only) |
| Convergent Validity (Known Groups)<br>Risk of a neurodevelopmental condition | 4000 | RBSK Scores | Area Under Curve (AUC) | Motor > 0.70*<br>Cognition > 0.70*<br>Social > 0.70*<br>(Total > 0.70)* |
| Convergent Validity (Known Groups)<br>Stunting/malnutrition | 4000 | Weight for age z-score (WAZ)<br>Height for age z-score (HAZ)<br>Weight for height z-score (WHZ) | Area Under Curve (AUC) | Motor > 0.70*<br>Cognition > 0.70*<br>Social > 0.70*<br>(Total > 0.70)* |
| Convergent Validity Household SES | 4000 | DHS Wealth Index | Pearson's correlation | R<sub>p</sub> > 0.20* |
| Convergent Validity Caregiver Education | 4000 | Highest level of school attended | Spearman's correlation | R<sub>s</sub> > 0.20* |
| Convergent Validity Caregiver depression | 4000 | PHQ-9 | Pearson's correlation | R<sub>p</sub> > 0.00* |
| Convergent Validity Participation | 2000 | Picture My Participation (PmP) | Pearson's correlation | R<sub>p</sub> > 0.00* |
| Convergent Validity Exposure to violence/ conflict; neglect/ abuse | 4000 | Childhood Psychosocial Adversity Scale (CPAS) | Pearson's correlation | R<sub>p</sub> > 0.00* |
| Convergent Validity Home Stimulation | 4000 | Family Care Indicators (FCI) | Pearson's correlation | R<sub>p</sub> > 0.20* |
| Convergent Validity Parent-child relationship | 4000 | Mother's object relations scale (MORS) (0-3yo)<br>Child Parent Relationship Scale (CPRS) (3-6yo) | Pearson's correlation | R<sub>p</sub> > 0.00* |
| Convergent Validity Social attention | 1000 | Various Neural/eye-tracking measures of development | Pearson's correlation | R<sub>p</sub> > 0.00* |
| Convergent Validity Exposure to stress | 200 | Hair cortisol | Pearson's correlation | R<sub>p</sub> > 0.00* |
| Responsiveness | 144 | STREAM<br>GMDS,<br>HAZ, WAZ, WHZ | Pearson's correlation of change scores between T1-T2 (18m) | R<sub>p</sub> > 0.20* |

\*Lower Two-Sided 95%CI > specified minimum

### Machine-learning

For domain score generation, a subset of features from the STREAM platform (similar to those described in the previous paragraph, on the construct-focused method) will be used, and optimisation will be performed against the target subset of the GMDS domain scores, which will serve as supervised learning labels. Our methodology will be validated by comparing the model-generated GMDS scores with the obtained GMDS scores on data in a verification set on which the machine-learning model has not been trained. Furthermore, we will scaffold the construct-focused approach of domain score generation with the machine-learning approach. Specifically, a parametric learning approach will be employed to generate scores that are correlated with seen data, thereby paving the way for future generation of construct-like scores for unseen data.

### Subgroup analyses

All relevant statistics will be reported by site, sex, and technology familiarity.

### Sample Size

Given the multiple goals of the study, multiple sample sizes have been estimated for specific purposes. A total sample size of N=4000 was found minimally sufficient to meet the various requirements of the study. This sample was broken down in multiple ways to minimise burden on individual participants and to collect the data required for analysis. For the factor model, Kline (2023) reports that a robust ratio of participants to estimated parameters is 20:1. Assuming our most complex cross-loaded model has 88 parameters (1 total score variance, 3 domain loadings, 3 correlations between domains, 27 task loadings, 27 task errors, 27 cross-loadings) this rule of thumb gives a required sample size of 1760. This is 90 less than our 1850 required to account for exclusions due to poor-quality responses. Note, all analyses below address non-inferiority to specified hypotheses (see Table 1) and, thus, use one-sided confidence intervals. For internal consistency, given a sample of 4000 and assuming an alpha of 0.85, the lower bound of our one-sided 95% confidence interval would lie at 0.84. For test-retest reliability, given a sample size of 150 and assuming a true reliability of 0.80, the lower bound of a one-sided 95% confidence interval lies at 0.75. For convergent validity evidenced through AUC, given a sample size of 4000, an expected 10% cases and assuming an AUC of 0.80, the lower bound of our one-sided 95% confidence interval lies at 0.73. For convergent validity evidenced through Pearson’s correlations, given a sample size of 4000, and expected correlations of 0.10 and 0.30, lower one-sided 95% CIs lie at 0.07 and 0.27 respectively. The same values hold true for Spearman’s correlation, to the two decimal places reported.

### Ethics and Dissemination

All STREAM components have been approved by local ethics review boards at each study site (India: Sangath Institutional Review Board; AIIMS Institute Ethics Committee; Indian Council of Medical Research - Health Ministry Screening Committee; Malawi: College of Medicine Research and Ethics Committee) and are carried out in accordance with Good Clinical Practice (GCP) standards.

### Benefits for families

Each family will receive informative handouts (Figure 3) covering topics such as nutrition, self-care, and child discipline methods, as well as a copy of their child’s growth chart following the anthropometric assessment. Additionally, each child will be given a gift (e.g., small toy, colouring pencils) and the family will receive monetary compensation for each visit (500 INR in India and 7700 MWK in Malawi) as a token of appreciation for their participation. Families will also be provided with refreshments during their visit.

### Referrals and support

The various STREAM assessments and questionnaires could potentially identify neurodevelopmental, health, or social difficulties (e.g., abuse or domestic violence) in children and/or their parents/caregivers. These cases may require support and referral to more specialist services via different pathways (Figure 3).

Children who are identified as having HAZ, WAZ, or WHZ scores of three standard deviations below the WHO child growth standards (WHO, 2006) will be referred to appropriate treatment services at both sites (e.g. Anganwadi Centres or nutrition clinics).

Children from the *Community* sample who are identified as having potential developmental delays (e.g. via RBSK, MDAT, GMDS, or INDTs) will receive appropriate referrals to specialist services such as tertiary hospitals (e.g. AIIMS in New Delhi, QECH in Malawi), government hospitals, paediatricians, and Early Intervention Centres (EICs). Parents of these children will be invited to engage in support sessions organised by the clinical psychologists on the research teams. These sessions aim to offer resources, guidance, and support related to child behaviour management at home, as well as suggestions for activities to promote environmental stimulation and support cognitive development. Additionally, parents will have the opportunity to engage in interactive sessions with other parents, where they can share experiences and provide suggestions on how to manage a child’s behaviour at home.

We will refer parents/caregivers who request support or advice in relation to questionnaire items measuring exposure to violence to non-governmental organisations (NGOs) for the protection of children and women and will provide them with details of support groups and helplines to contact. We will refer mothers who are identified as having maternal depression to psychiatrists, NGOs for mental health, and helplines.

### Dissemination

The STREAM project findings will be disseminated to participating families, healthcare professionals, policymakers, educators, and researchers, at local, national, and international levels. In Malawi, this might include the Ministry of Gender and Social Welfare, Ministry of Health, and organisations working with children with special needs. In India, this might include AIIMS, Maulana Azad Medical College, Ummeed Child Development Centre, Tamana, Action for Autism, ASHA and Anganwadi workers, and parents of children with and without developmental disabilities.

**Figure 3.**
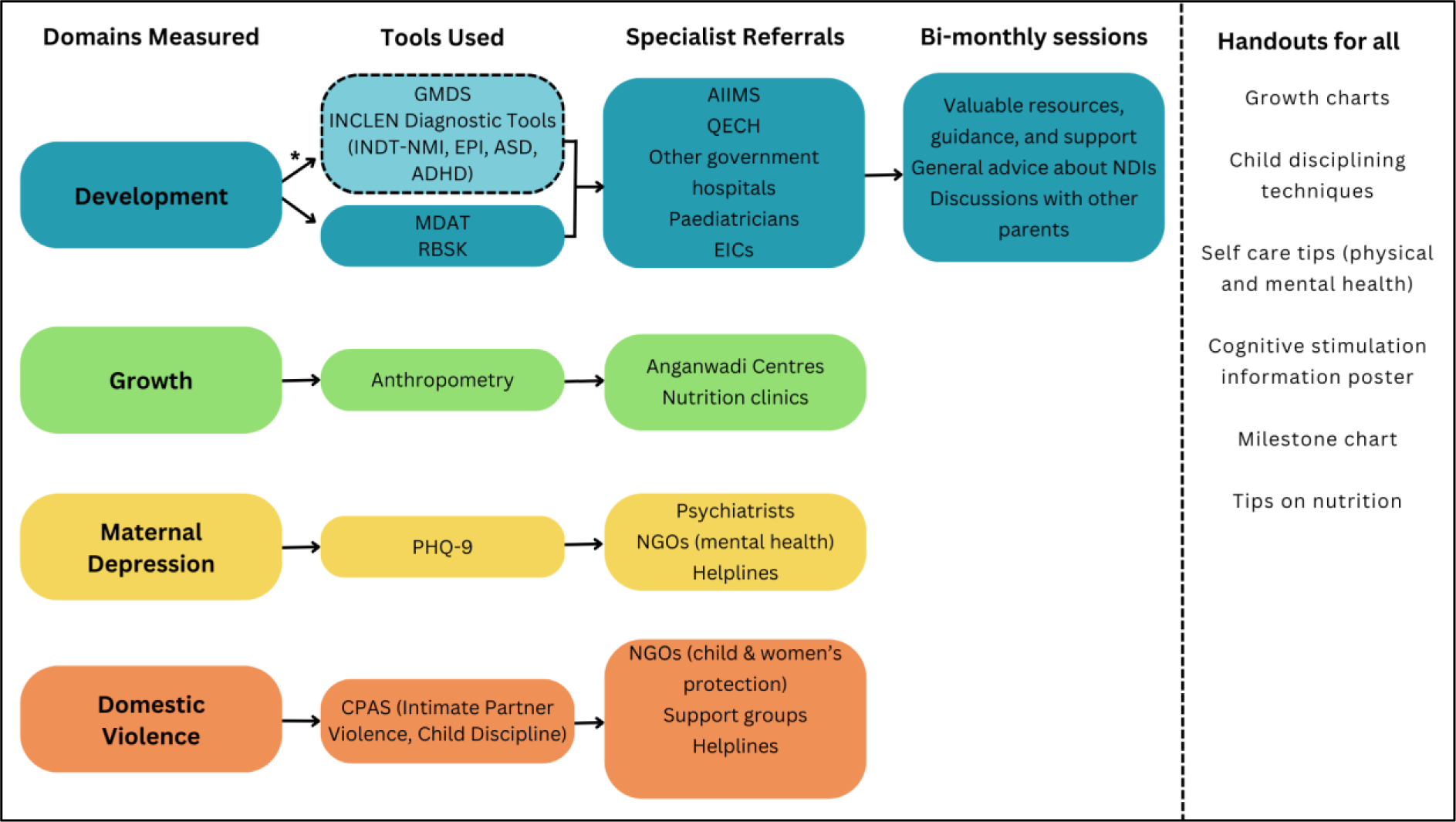
A flowchart illustrating the different referral and support pathways offered to families participating in STREAM. All families will be provided with various informative handouts. *This referral pathway is available only to children from the *Community* sample who participate in the Clinical Assessment.

## DISCUSSION

The STREAM project introduces a dimensional, scalable, tablet-based platform for non-specialist mediated assessment of neurodevelopmental status in three domains: social, cognitive, and motor. The adoption of a domain-based transdiagnostic approach is aligned with similar dimensional approaches in psychopathology (e.g. RDoC [Insel et al., 2010], HiTOP [Kotov et al., 2017]). This dimensional paradigm overcomes issues associated with categorical diagnostic approaches, such as comorbidity and phenotypic overlap between different neurodevelopmental conditions (e.g. Soke et al., 2018; Zhang et al., 2022).

Recent developmental assessment tools, such as the Global Scales for Early Development (GSED; Cavallera et al., 2023, McCray et al., 2023) and the Caregiver Reported Early Development Index (CREDI; McCoy et al., 2018) rely solely on parent-report and/or observation measures of development, which may be susceptible to issues such as recall bias, subjectivity, and inter-rater variability. STREAM aims to avoid these limitations by integrating parent-report and observation-based items with more direct measures of motor, social, and cognitive development. These direct measures include performance-based metrics derived from a child’s interaction with gamified tablet-based tasks, as well as automated video-based eye-tracking.

The MDAT is effective in identifying children aged 0-6 years with delays in social communication or motor functioning, while START and DEEP enable greater sensitivity and granularity in assessing social, motor, and cognitive processes in older children (2.5 to 6 years), who are able to effectively interact with a tablet device independently. Because STREAM integrates these three established tools into a single digital platform, it has the potential to assess motor, social, and cognitive functioning throughout the early developmental period from 0 to 6 years. This broad age-inclusivity aligns with the UN SDG of facilitating equal access to quality early-life care (UN SDGs, 2015).

The offline administration of tasks within the STREAM platform facilitates its use in remote settings. In contrast to paper-based instruments which are susceptible to recording and transcription errors, a tablet-based platform ensures more efficient and accurate data capture. Moreover, as the STREAM platform primarily relies on children’s engagement with tasks on a tablet-device, administration can be conducted by NSWs after training. These benefits will aid the future scalability of the STREAM platform and streamline its implementation in low-resource settings, as there is no requirement for any substantial investment in infrastructure or specialist human resources.

The STREAM platform has been designed to offer flexibility for future users. For example, stimuli and language for different contexts can be adapted through a user-friendly back-end content management system. The platform partitions light data (e.g. text, numeric) from heavy data (e.g. video recordings) within the back-end server, thereby optimising the efficiency of data access and download. Furthermore, from a scalability and software engineering perspective, the STREAM platform is set up as a docker container, simplifying the deployment of the back-end code on new servers for use in other projects. Moreover, the platform integrates various security features to ensure compliance with the UK Data Protection Act (2018) and the EU General Data Protection Regulation. These features safeguard the security of the data whether stored on the tablet-device, back-end server, or during transit between the two. This in-built flexibility and security further supports the scalability of the STREAM platform for future use in different settings, and through different research, clinical, or public health groups.

The validation of STREAM incorporates several design and methodological factors that adhere to the strict protocols necessary for validating an international measure of child development, such as those recommended in the COSMIN checklist (Mokkink et al., 2010). The study will test the psychometric properties of the STREAM platform in several ways, including internal and external reliability, concurrent and convergent validity, and sensitivity to change. The convergent measures capture constructs associated with child development using a diverse range of approaches, including self-report, biological (cortisol), and neural (EEG) measures. Furthermore, data collection is being undertaken in two low-resource settings which vary in terms of language, culture, socioeconomic status, and medical infrastructure, thus allowing some assessment of generalisability across diverse settings.

An aim of the STREAM project is to develop preliminary reference curves for both countries to index developmental norms analogous to those used for anthropometric outcomes (WHO, 2006). These reference curves will provide a valuable resource to facilitate the early identification of children facing challenges in any of the three developmental domains. It is important to note, however, that the reference curves generated within this project may have limited applicability beyond the contexts in which these data were collected. Data collection in other settings will be important to expand the validity and reliability of the data generated from the contexts used in the present study.

## Conclusion

This project will test the validity and reliability of the STREAM tablet-based platform in assessing social, cognitive, and motor development in children aged 0 to 6 years. Once validated, its implementation across low-resource settings has the potential to narrow the detection gap in identifying children who may benefit from additional support in certain developmental domains, enabling timely interventions and supporting children in achieving their full developmental potential.

## Data Availability

Not applicable

## Footnotes

1 The criterion of being ineligible for participation if currently enrolled in another research study or trial is relaxed for the *Enriched* sample in India because of its restrictive impact on recruitment numbers as most children presenting at the tertiary clinic will likely already be accessing care.

2 The models that describe only the separate domains will effectively be separate *factor analysis* models. Those that include a combined score will be *structural equation* models. These models will be fitted using the same set of procedures in the same statistical package. “Factor model” will henceforth be used to describe both types of models.

## Contributors

All authors contributed substantially to this work. EHW and NMT led the drafting of the manuscript. EHW, NMT, GM, MMCL, SB, DG, DM, TdB, GLE, LM, MKB, EJHJ, VP, SC, EM, GD, MG, and BC contributed to the study protocol. BC, MG, GD, EM, SC, VP, EJHJ, MKB, GL, MHJ, GM, DM, and SB led the conceptualisation of the study. MMCL, SB, DG, TdB, GLE, LM, VN, CN, RN, AB, AR, UK, NM, AS, IM, AG, SG, DM, and EM contributed to on-site implementation and drafting of site-or tool-specific details for the manuscript. GM and GL led the construct-focused analysis plan while SC and AG led the machine-learning analysis plan. All authors reviewed and edited the manuscript and read and approved the final version prior to submission.

## Funding

This work was supported by the Medical Research Council’s (MRC) Global Challenges Research Fund (GCRF), project reference MR/S036423/1.

## Competing Interests

None declared.

## Supplementary Materials

**Supplementary Materials 1:** Sex and age breakdown of the *Community* sample enrolled in PMA.

| Age Group | 0-3m | 3-6m | 6-9m | 9-12m | 1-1.5y | 1.5-2y | 2-2.5y | 2.5-3y | 3-3.5y | 3.5-4y | 4-4.5y | 4.5-5y | 5-5.5y | 5.5-6y | Total |
| --- | --- | --- | --- | --- | --- | --- | --- | --- | --- | --- | --- | --- | --- | --- | --- |
| Male | 77 | 77 | 77 | 77 | 154 | 154 | 154 | 154 | 154 | 154 | 154 | 154 | 155 | 155 | 1850 |
| Female | 77 | 77 | 77 | 77 | 154 | 154 | 154 | 154 | 154 | 154 | 154 | 154 | 155 | 155 | 1850 |
| **Total** | **154** | **154** | **154** | **154** | **308** | **308** | **308** | **308** | **308** | **308** | **308** | **308** | **310** | **310** | **3700** |

*\*To capture the rapid development occurring during the first year of life, 0 to 1 year olds will be stratified into 3-month intervals and all remaining years into 6-month intervals. The numbers specified within the table act as a general guide; it may not be feasible to recruit the exact numbers reported within each age bracket.*

**Supplementary Materials 2:** Sex and age breakdown of the *Enriched* Sample.

|  |  |  |  |  |
| --- | --- | --- | --- | --- |
| Targeted<br>NDIs<sup>+</sup> | GDD | GDD, ID,<br>ASC | ID, ASC,<br>ADHD |  |
| Age Group | 0-2y | 2-4y | 4-6y | **Total** |
| Male | 50 | 50 | 50 | **150** |
| Female | 50 | 50 | 50 | **150** |
| **Total** | **100** | **100** | **100** | **300** |

*\*The numbers specified within the table act as a general guide; it may not be feasible to recruit the exact numbers reported within each age bracket. <sup>+</sup>GDD: Global Developmental Delay; ID: Intellectual Disability; ASC: Autism Spectrum Conditions; ADHD: Attention Deficit Hyperactivity Disorder.*

**Supplementary Materials 3.**
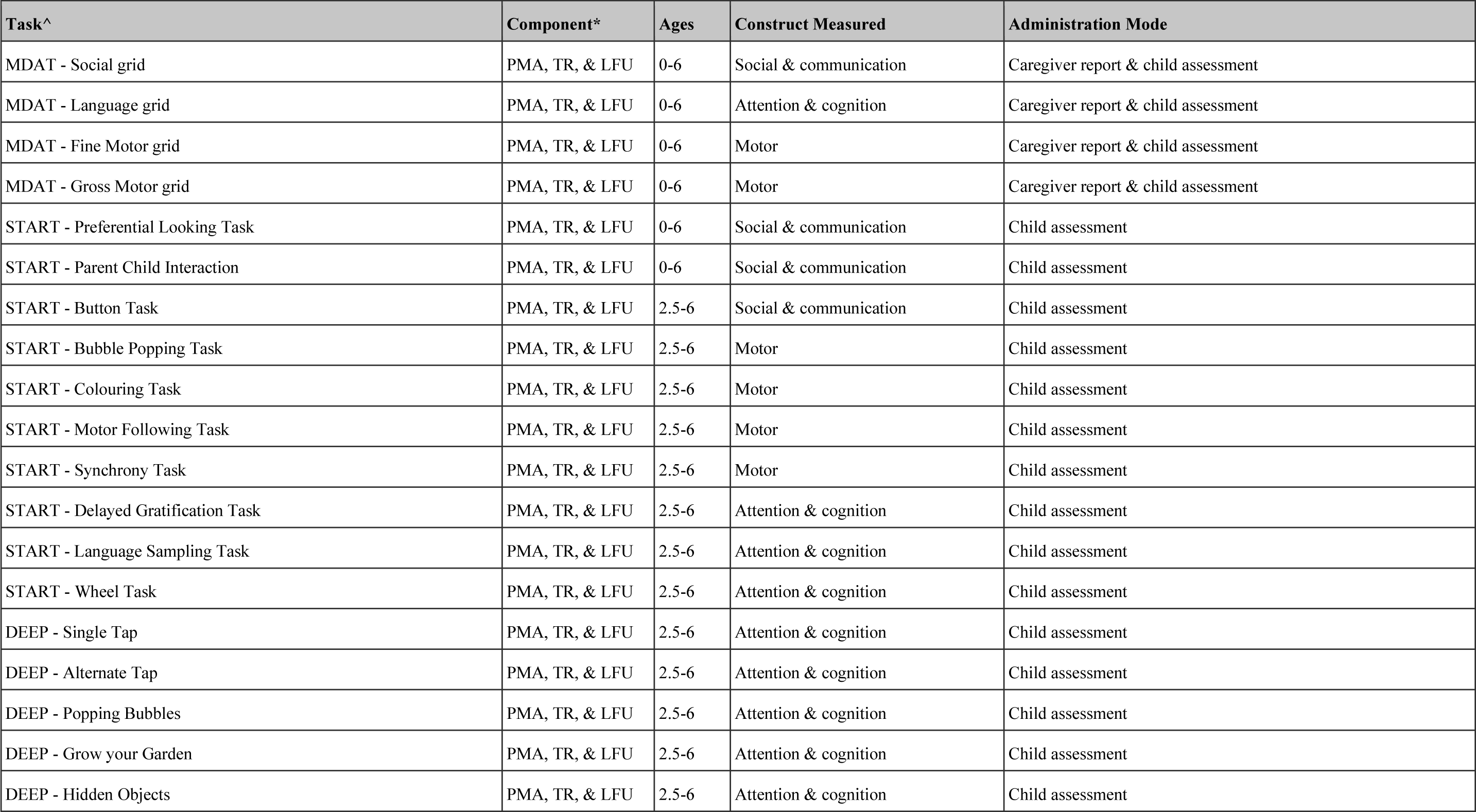
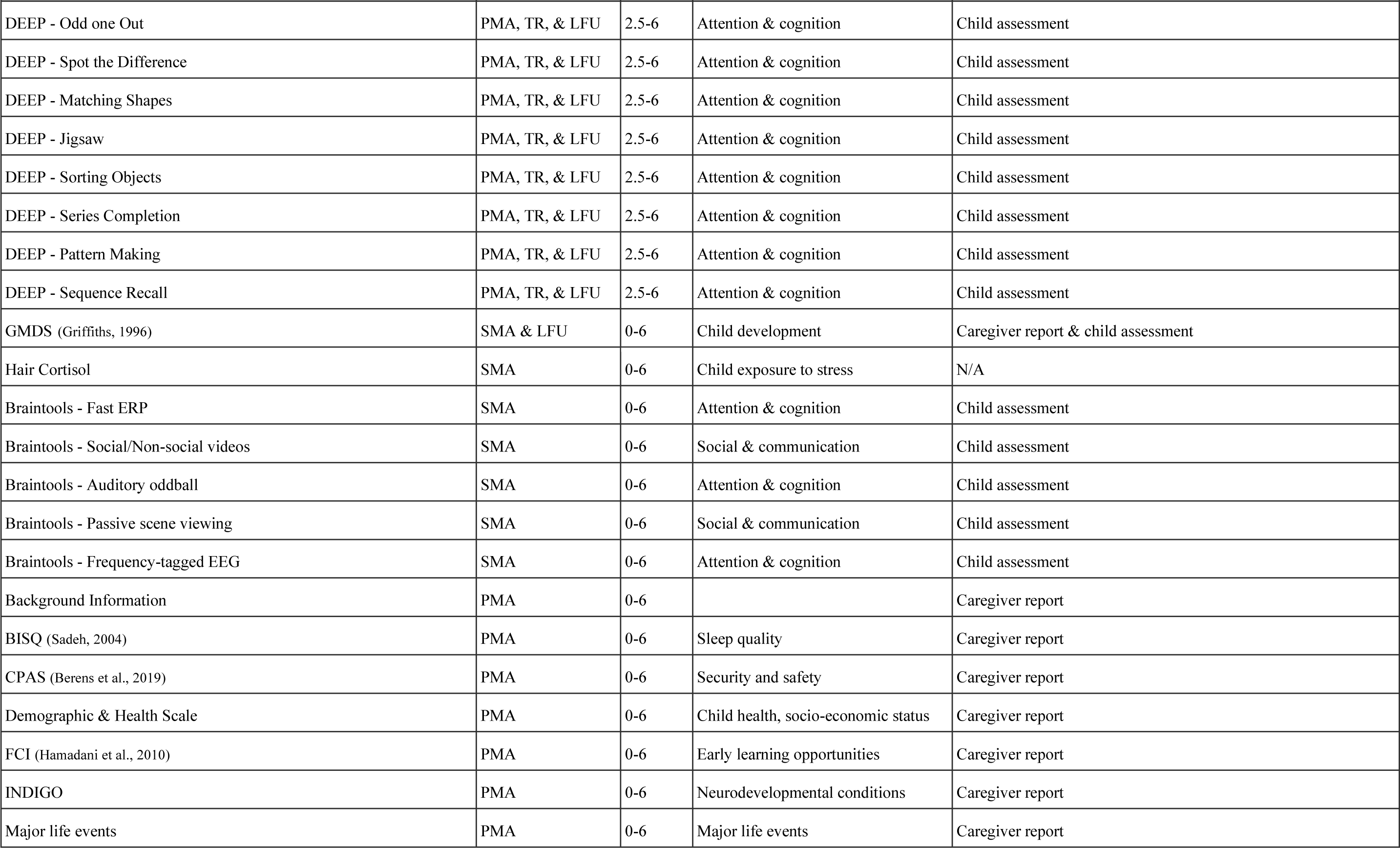
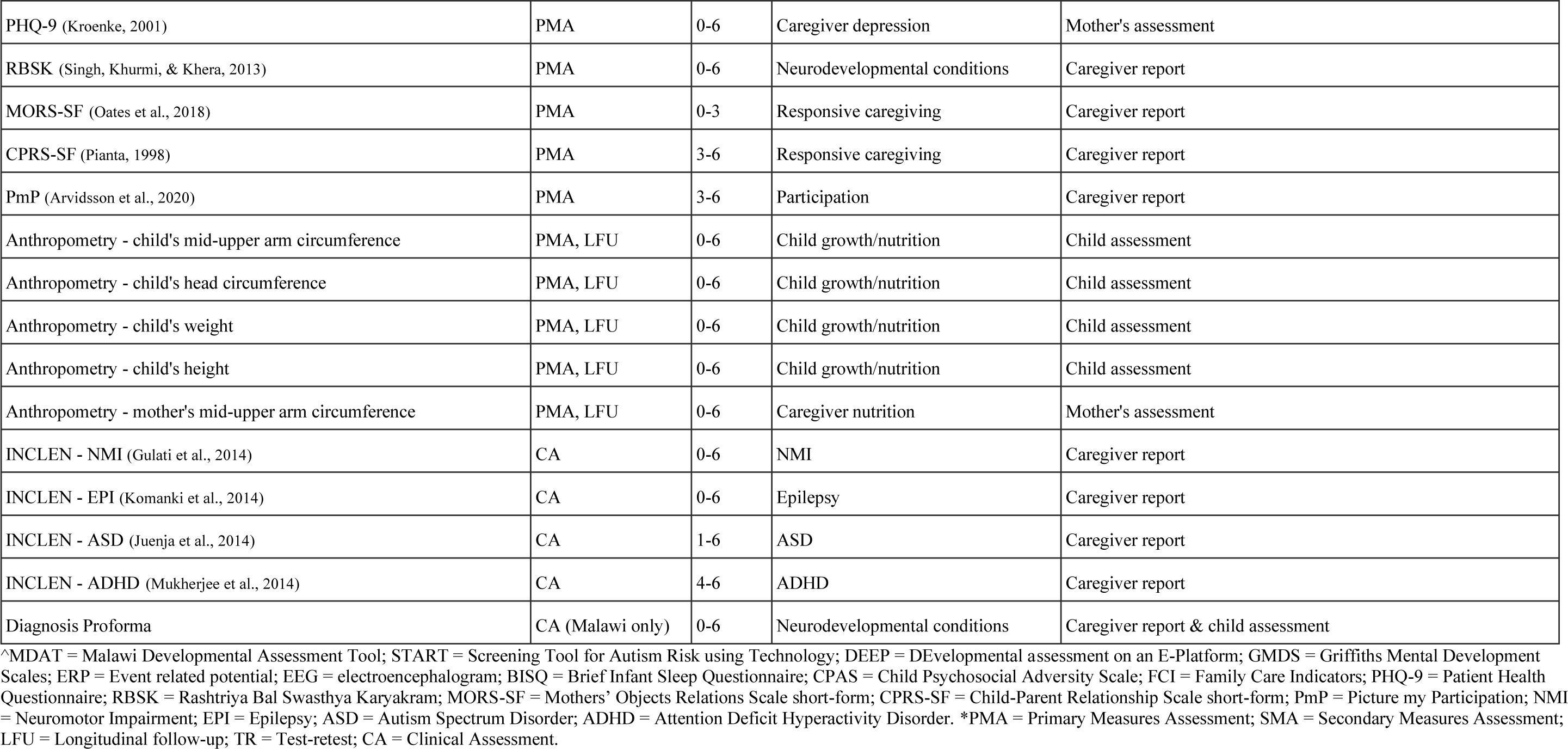
Overview of all measures included across each component of STREAM.

**Supplementary Materials 4:** Sex and age breakdowns for the Community sample enrolled in the SMA.

| Age Group | 0-3m | 3-6m | 6-9m | 9-12m | 1-1.5y | 1.5-2y | 2-2.5y | 2.5-3y | 3-3.5y | 3.5-4y | 4-4.5y | 4.5-5y | 5-5.5y | 5.5-6y | Total |
| --- | --- | --- | --- | --- | --- | --- | --- | --- | --- | --- | --- | --- | --- | --- | --- |
| Male | 21 | 21 | 21 | 21 | 41 | 42 | 41 | 42 | 41 | 42 | 41 | 42 | 42 | 42 | **500** |
| Female | 21 | 21 | 21 | 21 | 42 | 41 | 42 | 41 | 42 | 41 | 42 | 41 | 42 | 42 | **500** |
| **Total** | **42** | **42** | **42** | **42** | **83** | **83** | **83** | **83** | **83** | **83** | **83** | **83** | **84** | **84** | **1000** |

*\*To capture the rapid development occurring during the first year of life, 0-1 year olds will be stratified into 3-month intervals and all remaining years into 6-month intervals. The numbers specified within the table act as a general guide; it may not be feasible to recruit the exact numbers reported within each age bracket.*

**Supplementary Materials 5:** Clinical proforma.

CLINICAL EVALUATION
|  |
| --- |
| **Child ID:** |
| **Date of Assessment:** |
| **Child's age:** |
| **Exam by:** |

| Neurological Exam form |  |
| --- | --- |
| **1. Head abnormalities** | No abnormality ☐<br>Abnormality in shape (as seen in the pictures) ☐ |
| **2. Eye abnormalities** | ☐Yes ☐No |
| **3. Ear abnormalities** | ☐Yes ☐No |
| **4. Facial dysmorphism** | ☐Yes ☐No |
| **5. Upper or lower limb abnormalities** | ☐Yes ☐No |
| **6. Hip dislocation** | ☐Yes ☐No |
| **7. Scoliosis** | ☐Yes ☐No |
| **8. Neural tube defects (spina bifida, meningocele)** | ☐Yes ☐No |
| **9. Any features of a cleft lip or palate?** | ☐Yes ☐No |
| **10. Any other syndromic features** | ☐Yes ☐No<br>If yes, please describe _____ |
| **11. GMFCS level**<br>To show only for children above 2 years | **GMFCS level 1** ☐<br>**GMFCS level 2** ☐<br>**GMFCS level 3** ☐<br>**GMFCS level 4** ☐<br>**GMFCS level 5** ☐ |

| Diagnosis Pro-forma |  |
| --- | --- |
| **1. Do you think this child has a disability?** | Yes |
|  | No |
| **If YES to q1** |  |
| **1.1. How serious would you say it is?** | Mild |
|  | Moderate |
|  | Severe |
| **2. Do you think this child has a disability in any of the following areas? (Answer yes to more than one if you like)** | Motor skills |
|  | Cognition/learning |
|  | Hearing |
|  | Vision |
|  | Communication |
|  | Socio-Emotional difficulties (e.g., severe depression or anxiety) |
|  | Severe behavioral difficulties |
|  | Epilepsy |
| **3. In your opinion as a clinician, do you think the child is suffering from any of the following neurodisabilities? (Answer yes to more than one if you like)** |  |
| **3.1. Cerebral Palsy** | Yes |
|  | No |
|  | Not able to assess |
| **3.2. Neuromuscular condition** | Yes |
|  | No |
|  | Not able to assess |
| **3.3. Arthrogryposis** | Yes |
|  | No |
|  | Not able to assess |
| **3.4. Spina bifida** | Yes |
|  | No |
|  | Not able to assess |
| **3.5. Hydrocephalus** | Yes |
|  | No |
|  | Not able to assess |
| **3.6. Hearing impairment** | Yes, Unable to specify level |
|  | Yes, Moderate (41-70db) |
|  | Yes, Severe (71-90dB) |
|  | Yes, Profound (91+dB) |
|  | Not able to assess |
| **3.7. Visual impairment** | Yes, Unable to specify level |
|  | Yes, Moderate – visual acuity of <6/60 in better eye when best corrected |
|  | Yes, Severe – visual acuity of <3/60 in better eye when best corrected |
|  | Not able to assess |
| **3.8. Global Developmental Delay (>2SD in all areas of development or half developmental age expected)** | Yes |
|  | No |
|  | Not able to assess |
| **3.9. Concerns about Autistic spectrum disorder** | Yes |
|  | No |
|  | Not able to assess |
| **3.10. Congenital developmental disorder/syndrome** | Yes |
|  | No |
|  | Not able to assess |
| **3.11. Epilepsy** | Yes |
|  | No |
|  | Not able to assess |
| **3.12. ADHD** | Yes |
|  | No |
|  | Not able to assess |
| **4. In your opinion as a clinician, what's your final diagnosis for the child?** | [Free text] |
| **4.1. Please, add brief explanation justifying your decision** | [Free text] |

## Notes

### Competing Interest Statement

The authors have declared no competing interest.

### Author Declarations

Sangath Institutional Review Board gave ethical approval for this work. All India Institute of Medical Sciences Ethics Committee gave ethical approval for this work. Indian Council of Medical Research - Health Ministry Screening Committee gave ethical approval for this work. College of Medicine Research and Ethics Committee of University of Malawi gave ethical approval for this work.

